# Retention in paediatric and adolescent HIV care among ART initiation cohorts in Uganda, 2020–2023

**DOI:** 10.64898/2026.10.01.26364483

**Authors:** Kyomugisha Denise Aman, Richard Migisha, Ivan Lukabwe, Nasif Matovu, Anibare Nuwamanya, Vianney John Kigongo, Justine Wobusobozi, Winfred Nakaweesi, Anne Loy Alupo, Aminah Namwabira, Fatumah Nalubega, Lilian Bulage, Benon Kwesiga, Alex Riolexus Ario, Eleanor Namusoke Magongo

**Author notes:** Corresponding Author (KDA).

## Abstract

**Background:** Sustained retention in HIV care is essential for maintaining antiretroviral therapy (ART) and achieving viral suppression. We described national and regional retention in HIV care among paediatric and adolescent ART initiation cohorts in Uganda from 2020–2023.

**Methods:** We analyzed aggregated paediatric (aged 0-9 years) and adolescent (aged 10-19 years) ART cohort data reported through the District Health Information Software. We assessed 16 consecutive quarterly ART initiation cohorts per age group, from Quarter 1 2020 to Quarter 4 2023, at 6 (short-term), 12 (intermediate), and 24 (long-term) months after ART initiation. Each cohort comprised clients newly initiated on ART in a specific calendar quarter. Retention was defined as the proportion of clients alive and on ART within the net current cohort (original cohort plus transfer-ins minus transfer-outs). We described retention nationally and by region for each cohort and follow-up time point. We summarized the proportion of cohorts with retention ≥95% by region as a descriptive measure of high retention.

**Results:** Among paediatric cohorts, median retention was 84% (IQR: 82–85%) at 6 months, 81% (IQR: 78–83%) at 12 months, and 78% (IQR: 75–80%) at 24 months. Only one national paediatric cohort achieved ≥95% retention at 6 months and one at 12 months; none attained this target at 24 months. The proportion of paediatric cohorts with regional retention ≥95% was highest in Karamoja at short-term follow-up (31%), in Kigezi at intermediate follow-up (31%), and in Lango at long-term follow-up (13%).

Among adolescent cohorts, median retention was 82% (IQR: 80–85%) at 6 months, 82% (IQR: 81–85%) at 12 months, and 76% (IQR: 74–78%) at 24 months. No national adolescent cohort achieved ≥95% retention. The proportion of adolescent cohorts with regional retention ≥95% was highest in West Nile at 6 months (19%) and 12 months (31%), and in Karamoja at 24 months (25%).

**Conclusion:** Retention among paediatric and adolescent ART cohorts declined with longer follow-up and varied across regions. Strengthening age-appropriate support beyond the first year of ART and improving routine cohort-data quality could support more targeted programme responses.

## BACKGROUND

Retention in HIV care refers to the continued engagement of people living with HIV (PLHIV) in healthcare services after initiation of antiretroviral therapy (ART) (1). In practice, retention is used to describe a cohort of people living with HIV who are alive and receiving routine HIV care, including ART, at a specific time point after starting ART. As a program indicator, retention is defined as the proportion of PLHIV who are still alive and on ART at 6, 12 and 24 months after initiating ART (2). Retention is important because sustained engagement in care enables PLHIV to remain on ART thus contributing to the second UNAIDS 95-95-95 treatment target, which aims for 95% of all diagnosed PLHIV to receive sustained ART by 2030 (3–6).

Children and adolescents living with HIV require different forms of support to remain in HIV care. Younger children depend largely on caregivers for clinic attendance and treatment support, while adolescents may face challenges related to disclosure, stigma, school attendance, increasing independence and transition to adult care. These unique factors make the two age groups special interest groups for retention monitoring. Poor retention and interruptions in care may compromise viral suppression, treatment outcomes and may undermine the gains made after ART initiation (1,5,6). A systematic review of paediatric ART programmes found that up to 29% of children initiating ART were reported as dead or lost to follow-up by 12 months (7).

In Uganda, retention indicators are routinely reported through the national health information system (6,8). However, national evidence describing how retention differs between paediatric and adolescent ART initiation cohorts, across follow-up periods, and between regions remains limited. Retention in HIV care may vary geographically due to differences in health system performance, service accessibility, and population characteristics (7). Understanding geographic variation in retention outcomes is therefore important for area-specific program monitoring and planning, particularly for paediatric and adolescent populations who may face distinct barriers to sustained engagement in care (9,10). We described the variation in paediatric and adolescent retention in HIV care among ART initiation cohorts in Uganda during 2020–2023.

## METHODS

### Study design and setting

We conducted a retrospective descriptive analysis of routinely reported ART cohort data from health facilities providing HIV care in Uganda. ART cohort data are reported through the Ministry of Health national health information system. Clients initiating ART are grouped into quarterly initiation cohorts and subsequently followed using standardized cohort reports that capture treatment outcomes at predefined follow-up intervals.

Retention in HIV care is assessed at 6, 12, and 24 months after ART initiation, corresponding to follow-up quarters determined by the original initiation quarter. We considered 6 months as short-term, 12 months as intermediate, and 24 months as long-term follow-up time points.

### Study population

We included consecutive ART initiation cohorts of paediatric (aged 0–9 years) and adolescent (aged 10–19 years) data from quarter 1 of 2020 to quarter 4 of 2023. Paediatric cohorts comprised children aged 0–9 years, whereas adolescent cohorts comprised individuals aged 10–19 years. We defined these age groups at ART initiation.

### Data sources and study variables

We extracted routinely reported ART cohort data from the Uganda Ministry of Health District Health Information Software version 2 (DHIS2). We disaggregated data by ART initiation quarter, age group, follow-up time point, and region. For each cohort and follow-up time point, the variables used in this analysis were the number of cohort members who were alive and on ART and those in the net current cohort. The net current cohort comprised the original ART initiation cohort, adjusted for transfer-ins and transfer-outs.

### Outcome definition

The primary outcome was retention percentage, defined as the proportion of clients who were alive and on ART within the net current cohort during the relevant calendar quarter.

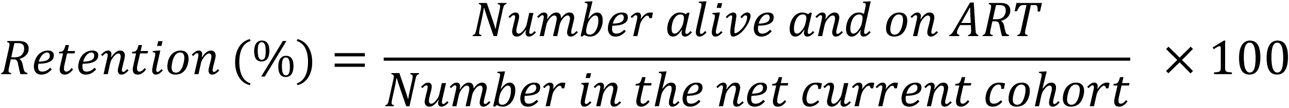

Nationally, cohort-specific retention percentages were summarized across the 16 quarterly ART initiation cohorts for each age group and follow-up time point. Regionally, cohort-specific retention percentages were similarly summarized within each region for each age group and follow-up time point.

We assessed achievement of the ≥95% retention target as a secondary outcome. For each region and follow-up time point, we determined the number and proportion of quarterly ART initiation cohorts with retention at or above 95%.

### Study size

We included all available ART initiation cohorts in DHIS2 that had completed sufficient follow-up time to allow assessment at 6, 12, and 24 months after ART initiation.

### Data analysis

Data were analyzed using R statistical computing software version 4.2.3. Missing retention observations were excluded from analysis. Implausible or unusually high or low retention values were retained without modification.

For each region, age group and follow-up time point, cohort-specific retention percentages were summarized across the 16 quarterly ART initiation cohorts using the median and interquartile range (IQR), along with the minimum and maximum values. National retention was visualized using bar charts showing age-specific cohort retention against the ≥95% retention target by time point. Regional variation in median retention was visualized using heatmaps and choropleth maps generated in Quantum Geographic Information System (QGIS) software. As a secondary analysis, we assessed regional consistency in attaining the ≥95% retention target. For each region, age group and follow-up time point, we calculated the number and proportion of quarterly ART initiation cohorts that attained the target.

## RESULTS

### National retention percentages

For paediatric cohorts, 6-month retention ranged from 77% to 95%, with only the Q4 2023 cohort attaining the ≥95% target. At 12 months, retention ranged from 64% to 109%, with only the Q2 2020 cohort exceeding the target. At 24 months, retention ranged from approximately 39% to 82%, and none of the cohorts met the target. Q3 2023 cohort had the lowest paediatric retention, at approximately 39% (Figure 1).

**Fig 1.**
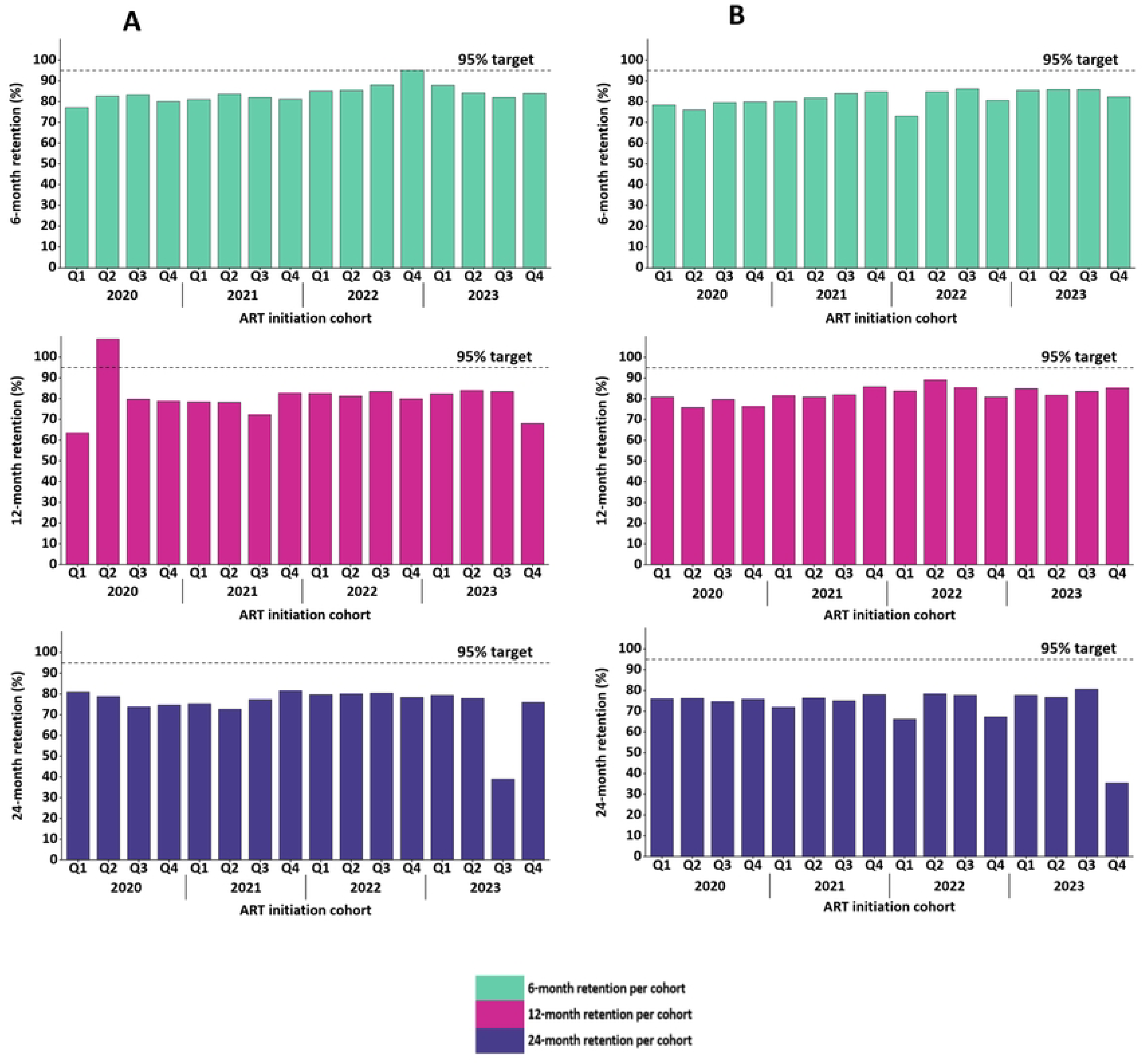
National paediatric (A) and adolescent (B) retention percentages at 6 (short-term), 12 (intermediate) and 24 months (long-term) follow-up time points among individual 2020–2023 ART initiation cohorts in Uganda.

For adolescent cohorts, 6-month retention ranged from 73% to 86%; 12-month retention ranged from approximately 76% to 89%; and 24-month retention ranged from 36% to 81%, and none of the adolescent cohorts met the target at any time point. Q4 2023 cohort had the lowest adolescent retention, at approximately 36% (Fig 1).

### National median retention percentages

Among both paediatric and adolescent cohorts, median retention percentages were highest at short-term follow-up (77–95 and 73–86, respectively) and lowest at long-term follow-up (39–82 and 36–81, respectively) (Table 1).

**Table 1.** National median retention percentages across the 16 quarterly ART cohorts by age group and follow-up time point in Uganda, 2020–2023.

| Retention follow-up time point | Median retention, % | (IQR) | Range % |
| --- | --- | --- | --- |
| **Paediatric retention** |  |  |  |
| Short-term | 84 | (82–85) | 77–95 |
| Intermediate | 81 | (78–83) | 64–109 |
| Long-term | 78 | (75–80) | 39–82 |
| **Adolescent retention** |  |  |  |
| Short-term | 82 | (80–85) | 73–86 |
| Intermediate | 82 | (81–85) | 76–89 |
| Long-term | 76 | (74–78) | 36–81 |

### Regional median retention percentages

Median paediatric retention varied across regions and follow-up time points. At short-term follow-up, most regions had median retention in the 80–94% category. At intermediate follow-up, regional median retention was more varied, with most regions falling within the 65–79% and 80–94% categories. At long-term follow-up, a greater proportion of regions had median retention in the 65–79% category. No region had a median paediatric retention of ≥95% at any follow-up time point (Fig 2A and Fig 2B).

**Fig 2A.**
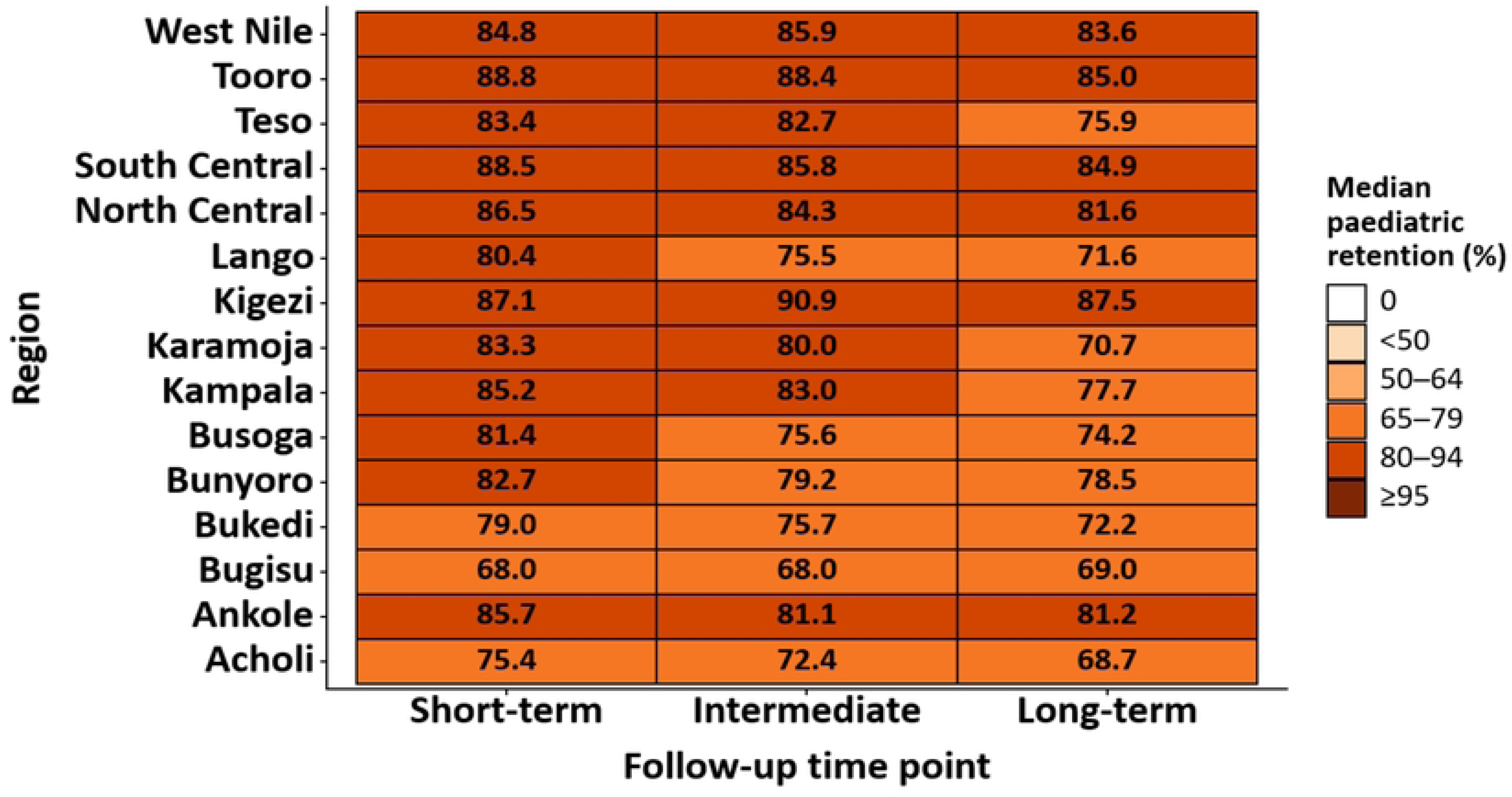
Heatmap of median paediatric retention by region at short-term, intermediate and long-term follow-up among 2020–2023 ART initiation cohorts in Uganda.

**Fig 2B.**
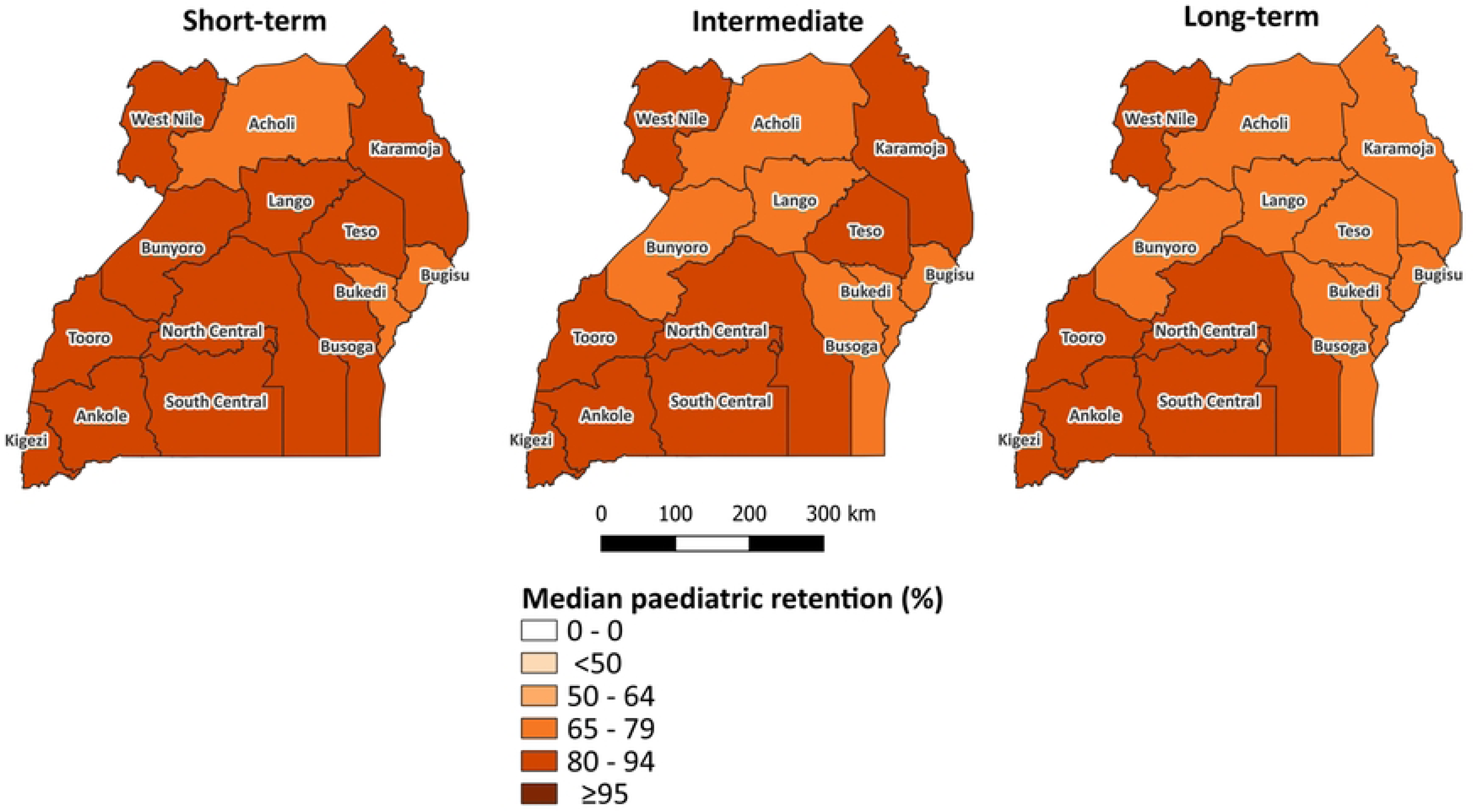
Regional distribution of median paedoatric retention by region at short-term, intermediate and long-term follow-up among 2020–2023 ART initiation cohorts in Uganda.

Median adolescent retention varied across regions and follow-up intervals. At short- and intermediate-term follow-up, regional median retention was generally higher, with more regions in the 80–94% category. At long-term follow-up, more regions had median retention in the 65–79% category. No region had a median adolescent retention of ≥95% at any follow-up time point (Fig 3A and Fig 3B).

**Fig 3A.**
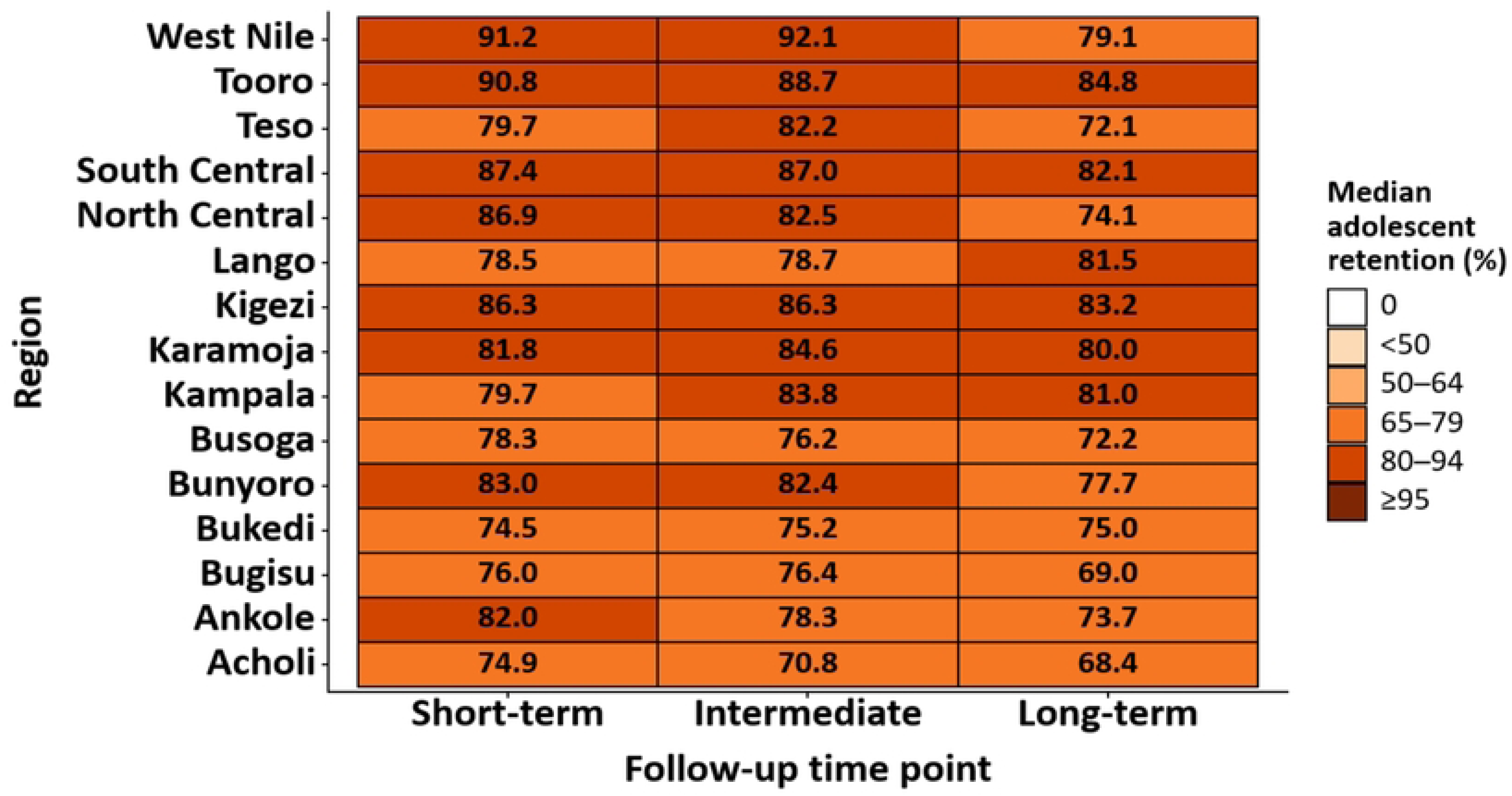
Heatmap of median adolescent retention by region at short-term, intermediate and long-term follow-up among 2020–2023 ART initiation cohorts in Uganda.

**Fig 3B.**
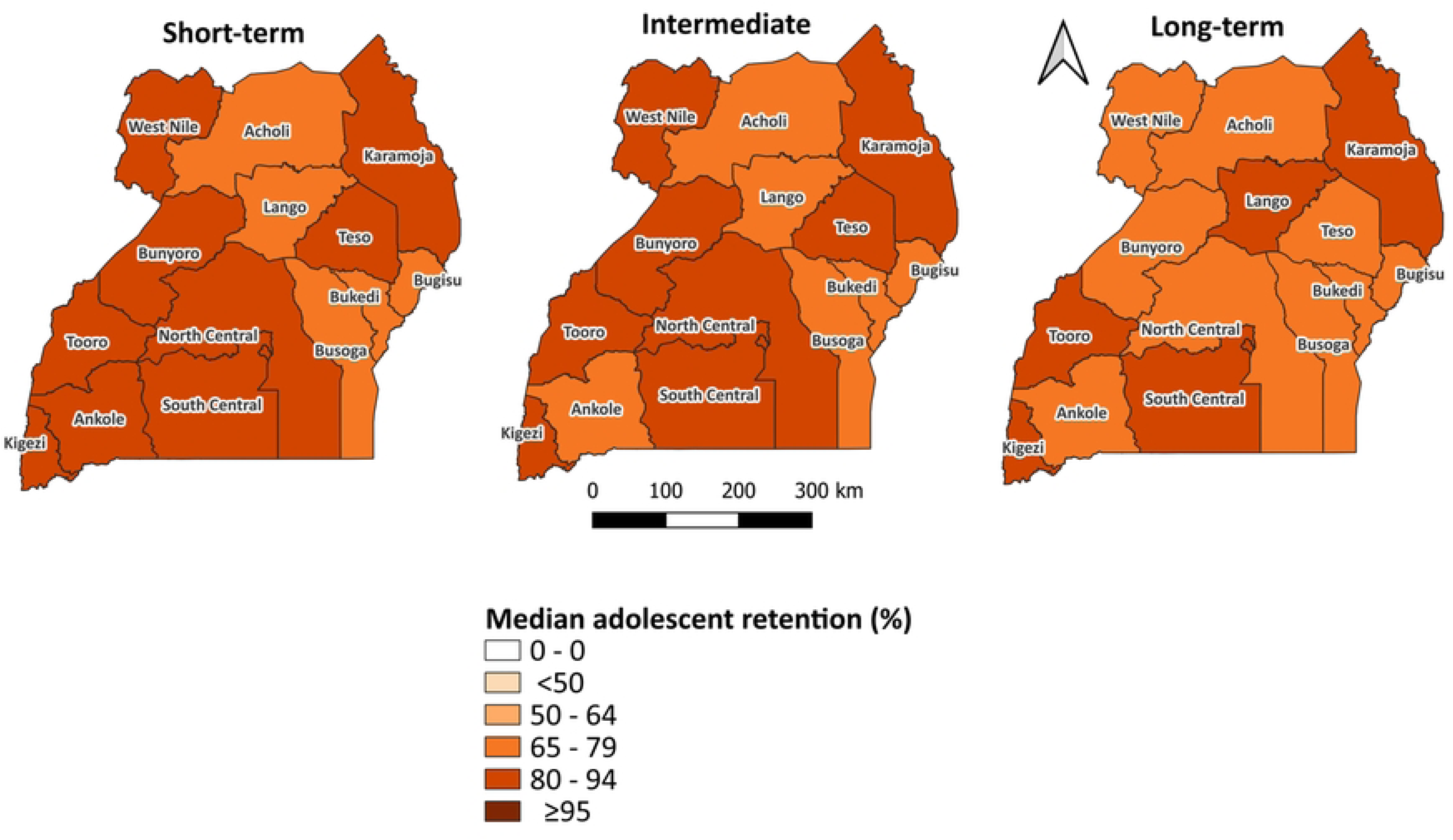
Regional distribution of median adolescent retention by region at short-term, intermediate and long-term follow-up among 2020–2023 ART initiation cohorts in Uganda.

### Regional consistency in attaining the ≥95% retention target

Attainment of the ≥95% retention target was rare and varied by region, age group, and follow-up time point. Among paediatric cohorts, Karamoja had the highest percentage attaining the target at short-term follow-up (5/16; 31%), while Kigezi led at intermediate follow-up with the same proportion. At long-term follow-up, success was lower across regions, with Lango having the highest proportion, (2/16; 13%). Bukedi, Bunyoro, Busoga, Kampala, North Central, and South Central had no pediatric cohorts reaching the target at any follow-up time point.

Among adolescent cohorts, West Nile had the highest percentage reaching the target at both short-term and intermediate follow-ups (3/16; 19%) and (5/16; 31%), respectively. At long-term follow-up, Karamoja had the highest proportion (4/15; 27%) meeting the target. Kampala and North Central had no adolescent cohorts achieving the ≥95% retention target at any follow-up point (Table 2 and Supplementary figure 1).

**Table 2:**
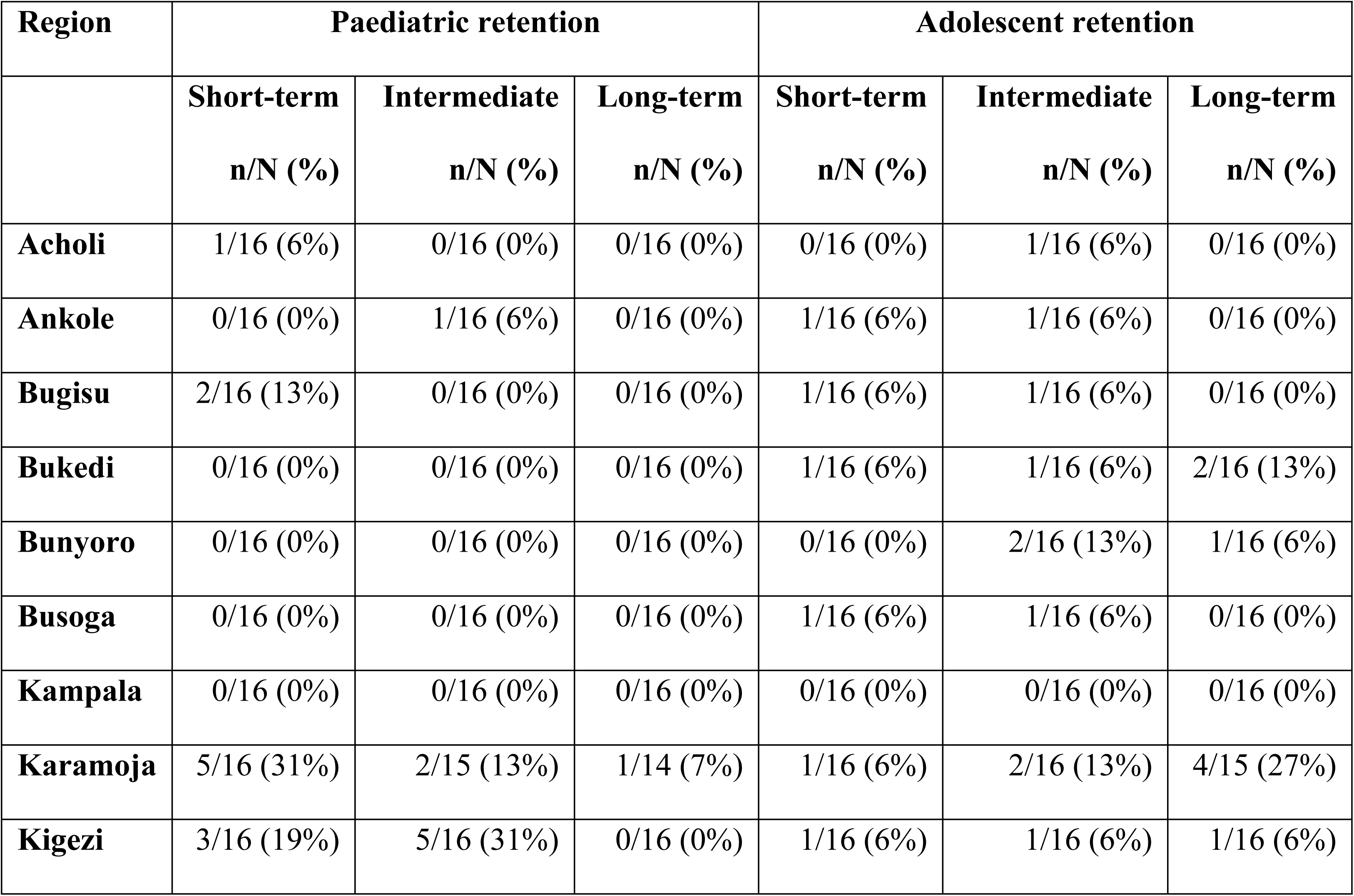
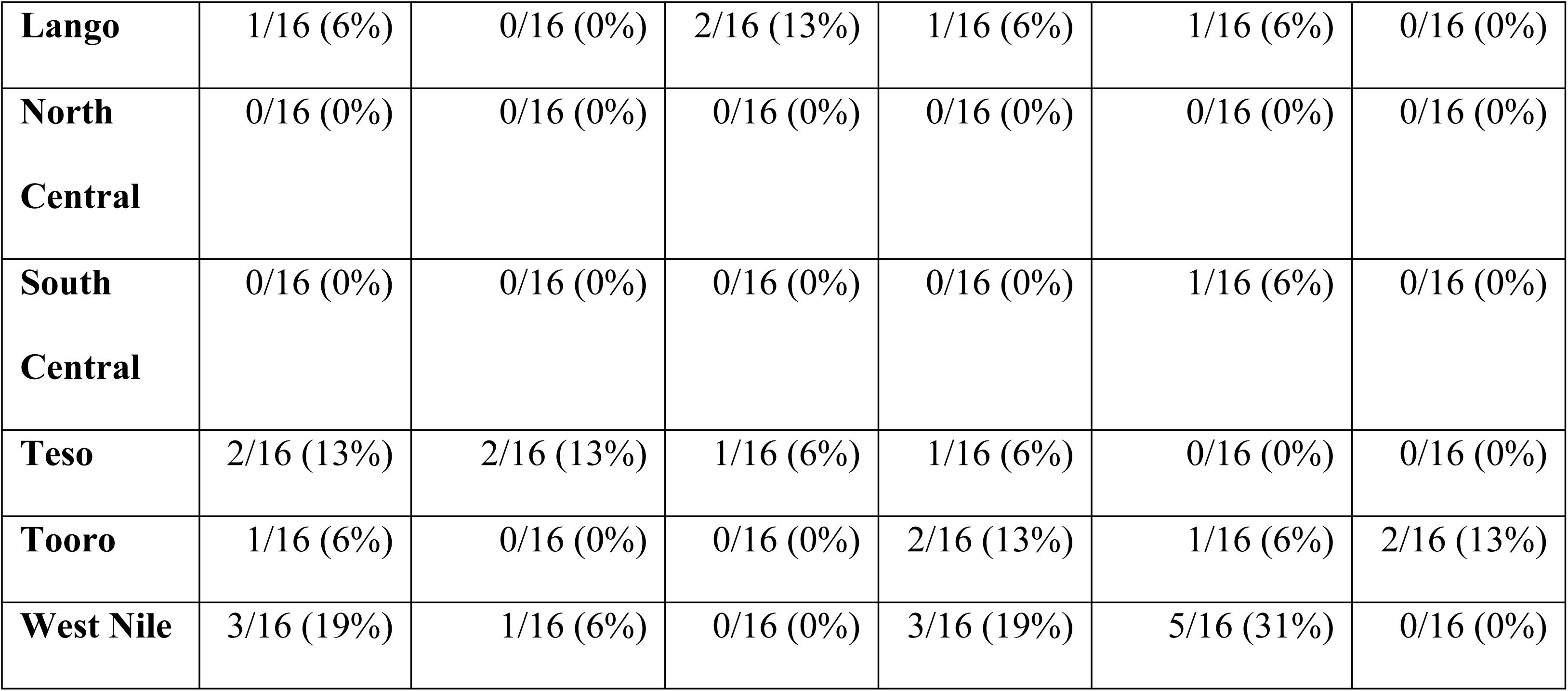
Regional consistency in attaining the ≥95% retention target among 2020–2023 paediatric and adolescent ART initiation cohorts in Uganda.

## DISCUSSION

Retention was higher at short-term (6-month) and intermediate (12-month) follow-up than at long-term (24-month) follow-up among both paediatric and adolescent ART initiation cohorts. Attainment of the ≥95% programme target was uncommon, with only two paediatric cohorts attaining the national target and no adolescent cohort attaining it at any follow-up time point. Regional median retention also varied by follow-up time point, with more regions having lower median retention at long-term than at short-term or intermediate follow up time points. Overall, the findings indicate that long-term retention is an important programme challenge for both age groups. HIV programmes should therefore give greater attention to supporting retention beyond the first year of treatment.

The median intermediate paediatric retention observed in this analysis was similar to estimates reported from other African ART programmes, although substantial variation was observed among the individual Ugandan cohorts. One multi-country cohort study reported paediatric retention of 80% at 12 months and 72% at 24 months in Kenya, Mozambique, Rwanda and Tanzania (10). A meta-analysis reported 12-month retention ranging from 71% to 95% across resource-limited settings (11). Although the median 12-month retention observed in Uganda was comparable with these reports, most individual cohorts remained below the ≥95% programme target. Therefore, comparable retention levels across settings should not be interpreted as indicative of adequate programme performance.

Median paediatric retention was highest at short-term and lower at long-term follow-up. Since younger children depend on caregivers for clinic attendance, medicine collection and treatment support, paediatric retention services need to consider the caregiver and the child together. Studies in Uganda have examined family-centred and community-based approaches to maintaining children and adolescents in care (12,13). In the Uganda Family Clinic Day trial, family-centred scheduling and health education improved adherence to clinic appointments, although it did not significantly improve six-month retention (13). These findings suggest that caregiver engagement and practical appointment support are still relevant components of paediatric HIV care, but further evidence is needed to identify which approaches improve long-term retention.

Median adolescent retention was similar at short-term and intermediate follow-up but was lower at long-term follow-up. No adolescent cohort attained the national ≥95% retention target, while regional target attainment occurred in selected cohorts and was most frequent at intermediate follow-up. However, attainment was not consistent across cohorts or regions. A Ugandan study similarly found that adolescent retention varied by treatment site and that older adolescents had a greater risk of attrition (14). Studies in Uganda have also identified school attendance, stigma and disclosure concerns as barriers to adolescent engagement in HIV care (15). Adolescent retention should therefore not be managed simply as an extension of paediatric or adult care. Reviews of transition in sub-Saharan Africa have highlighted the need for planned, youth-responsive transition arrangements between paediatric and adult HIV services (16). Appointment tracking, adolescent-focused clinic sessions and regular discussions with adolescents and caregivers improved retention in a quality-improvement project in western Uganda, although the findings came from one facility and should be interpreted cautiously (17).

Regional variation was another important finding. Median retention differed across regions and follow-up time points for both paediatric and adolescent cohorts, with more regions having lower median retention at long-term follow-up. Although no region had a median retention ≥95%, individual cohorts within some regions attained the target. However, target attainment was not consistent across cohorts or follow-up intervals. National summaries may therefore conceal both local strengths and areas requiring additional support. Studies using routine HIV programme data have also demonstrated substantial variation in outcomes between health care facilities (18). Uganda’s HIV programme should examine regions with comparatively higher retention and determine whether their service-delivery practices, programme implementation or data-quality factors contribute to the observed differences. This approach is supported by the Uganda positive-deviance study that identified practices associated with long-term retention within community ART delivery groups (19).

Regions with comparatively lower retention should undergo similar reviews to identify local service-delivery and data-quality gaps. A Ugandan quality improvement study showed that identifying site-specific causes of poor performance and testing locally feasible changes could improve adolescent retention (17). Programme support should therefore respond to locally identified gaps rather than relying on one uniform intervention.

A recent Ugandan programme review reported 87.8% retention among children and adolescents receiving multi-month ART refills (20). Although the study population and retention definition differed from those used in this analysis, the reported retention estimate was similar to the levels observed among several cohorts in our analysis and remained below the ≥95% programme target. Multi-month ART refills may address some barriers related to frequent clinic attendance, although additional strategies may be needed to support sustained retention. Reviews of differentiated service delivery have found generally favourable retention outcomes, but evidence specifically concerning adolescents remains limited (21,22).

A key strength of this study is its use of national routine programme data covering paediatric and adolescent ART initiation cohorts across Uganda. Assessing retention at six, 12 and 24 months allowed comparison of short-, intermediate and long-term retention. Summarizing actual retention percentages nationally and by region also allowed us to describe variation in retention.

The main limitation is that the study used aggregated DHIS2 data. As a result, we could not examine the patient, caregiver, clinical, facility or programme factors associated with retention. Secondly, regional estimates may conceal important differences between districts and healthcare facilities. Summarizing retention across quarterly cohorts using medians may mask variation between individual cohorts within the same region, although IQR and range were used to describe this variation. The broad age categories also prevented separate assessment of younger and older children and adolescents. In addition, unusually high or low retention values were retained as reported and may reflect programme performance, reporting errors or cohort reconciliation issues. These observations require data validation before they can be interpreted as programme performance.

### Public health and policy implications

Uganda’s HIV programme should place greater emphasis on retention beyond the first year of ART, given the lower retention observed at long-term follow-up in both age groups.

Routine programme reviews should include 24-month retention and compare retention with viral-load suppression to provide a fuller picture of treatment outcomes. Programme support should be tailored to age-specific and regional retention patterns, while routine data validation and cohort reconciliation should be strengthened to improve interpretation of estimates and identify areas requiring additional support.

## CONCLUSION

In conclusion, paediatric and adolescent retention was higher at short-term and intermediate follow-up than at long-term follow-up, with lower median retention at 24 months in both age groups. Retention also varied across regions and follow-up time points, while attainment of the ≥95% programme target was uncommon and inconsistent across cohorts. Uganda’s HIV programme should strengthen retention support beyond the first year of ART, tailor services to age-specific and regional retention patterns, and improve the quality and use of routine cohort data for programme decision-making.

## Data Availability

The datasets upon which our findings are based belong to the Uganda Public Health Fellowship Program. For confidentiality reasons, the datasets are not publicly available. The datasets can be availed upon reasonable request from the corresponding author with permission from the Uganda Public Health Fellowship Program. Request can be directed to.

## AUTHORS’ CONTRIBUTION

KDA conceived, designed, analyzed, interpreted the study and wrote the draft manuscript. KDA, NM, AN, VJK, JW, WN, ALA, AN and FN technically reviewed and edited the manuscript. IL, LB, BK, RM, ENM, ARA critically reviewed the manuscript for intellectual content. All authors reviewed and approved the final manuscript for publication.

## ETHICAL CONSIDERATIONS

This analysis utilized routinely generated aggregated surveillance data with no personal identifiers, abstracted from the District Health Information Software version 2 (DHIS2), which is managed by the Ugandan Ministry of Health. Access to DHIS2 data is granted by the Ministry of Health Data and Health Informatics Unit. Through this authorization, the Uganda National Institute of Public Health accessed and analysed this surveillance data to inform public health decision-making and HIV service delivery for children and adolescents, as well as obtained permission to disseminate the findings through scientific publications. We stored the abstracted dataset on password-protected computers and restricted access to only the investigation team. The Office of the Associate Director for Science at the US Centres for Disease Control and Prevention (CDC) Uganda reviewed the activity and determined that it did not constitute human subject research and that its primary purpose was public health practice and disease control. This activity was reviewed by the US CDC and conducted consistent with applicable federal laws and CDC policy namely; 45 C.F.R. part 46; 21 C.F.R. part 56; §42 U.S.C. §241(d); 5 U.S.C. §552a; 44 U.S.C. §3501 et seq.

## ACKNOWLEDGMENTS

The authors convey their gratitude to the Ministry of Health through the AIDS Control Program and Department of Health Informatics for the technical support provided.

## Supporting information

**S1 Fig.**
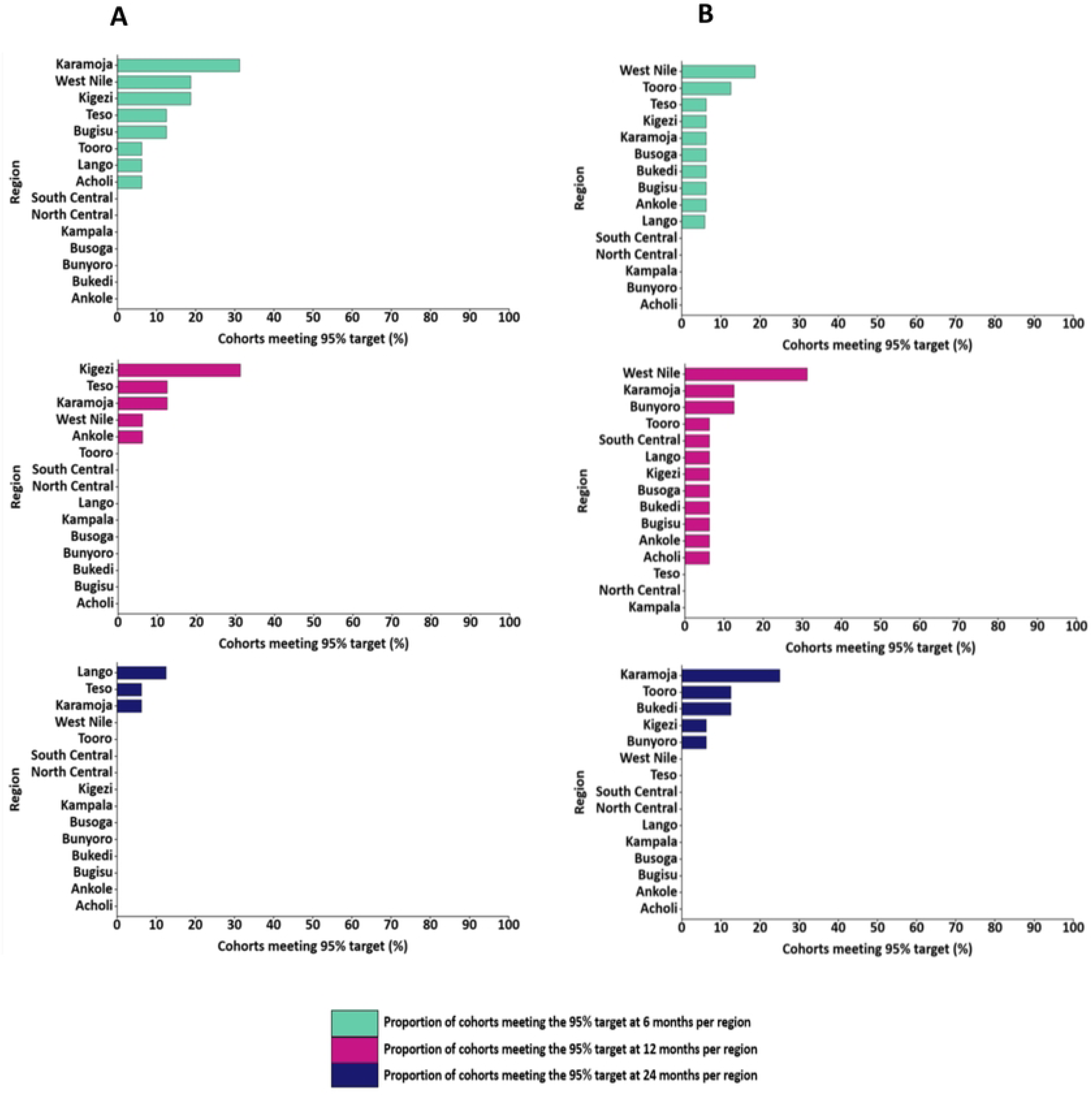
Regional consistency in attaining the ≥95% retention target among 2020-2023 paediatric and adolescent ART initiation cohorts in Uganda.

